# Simultaneous exposure to organophosphate and pyrethroid pesticides and effects on brain health in humans. A systematic review

**DOI:** 10.1101/2025.08.19.25334018

**Authors:** María Teresa Muñoz-Quezada, Rocío Hojas, Joaquín Toro, José Norambuena, Boris Lucero, Tristan Bekinschtein, Liliana Zúñiga, Cristian Valdés, Andrés Canales-Johnson, Ramón Castillo, Juan Pablo Gutiérrez-Jara, Benjamín Castillo, Natalia Landeros, Cynthia Carrasco, Jandy Adonis, Patricio Olguín

## Abstract

Organophosphate and pyrethroid pesticides are widely used, but their combined effects on brain health remain unclear. This study analyzed observational research from 2014 to 2024 on their simultaneous exposure and its association with neurobehavioral, neurodegenerative, and neurological disorders through a systematic review following PRISMA guidelines in PubMed and Web of Science. Studies assessing co-exposure with validated methods and reporting brain health outcomes were included, while reviews, animal studies, and those without simultaneous exposure data were excluded. Risk of bias in cohort studies was evaluated using ROBINS-E. Of the 34 studies included, nine were cohort studies, four with low risk of bias and four with concerns related to sample size or exposure assessment. Three linked co-exposure to attention deficits and cognitive decline in children. Evidence on neurodegenerative diseases and autism spectrum disorder was inconsistent due to conflicting results. Cross-sectional and case-control studies associated co-exposure with depression and anxiety but lacked follow-up data. Variability in exposure assessment reduced comparability. Research remains limited, indicating possible neurodevelopmental effects and the need for standardized methodologies to refine risk assessment.

## Introduction

Pesticide exposure represents a significant public health concern due to the widespread use of these chemicals in agriculture, pest control, and residential settings (Dereumeaux et al., 2019) While pesticides are primarily utilized to enhance agricultural productivity and manage vector-borne diseases, their unintended adverse effects on human health remain a critical issue (World Health Organization, 2019).

Insecticidal pesticides, including organophosphates (OP) and pyrethroids (PYR) pesticides, are among the most commonly employed agents worldwide due to their effectiveness in controlling pests (Figure 1). However, their neurotoxic potential, particularly under conditions of chronic or simultaneous exposure, raises serious health concerns (Hirano et al., 2023). OP function by inhibiting acetylcholinesterase, an enzyme essential for breaking down acetylcholine in the nervous system. This inhibition leads to acetylcholine accumulation, resulting in neurological symptoms that range from mild (e.g., headaches and dizziness) to severe (e.g., seizures, neurobehavioral impairments, and long-term neurodegenerative disorders) (Ganie et al., 2022; Masood et al., 2024).

**Figure 1.**
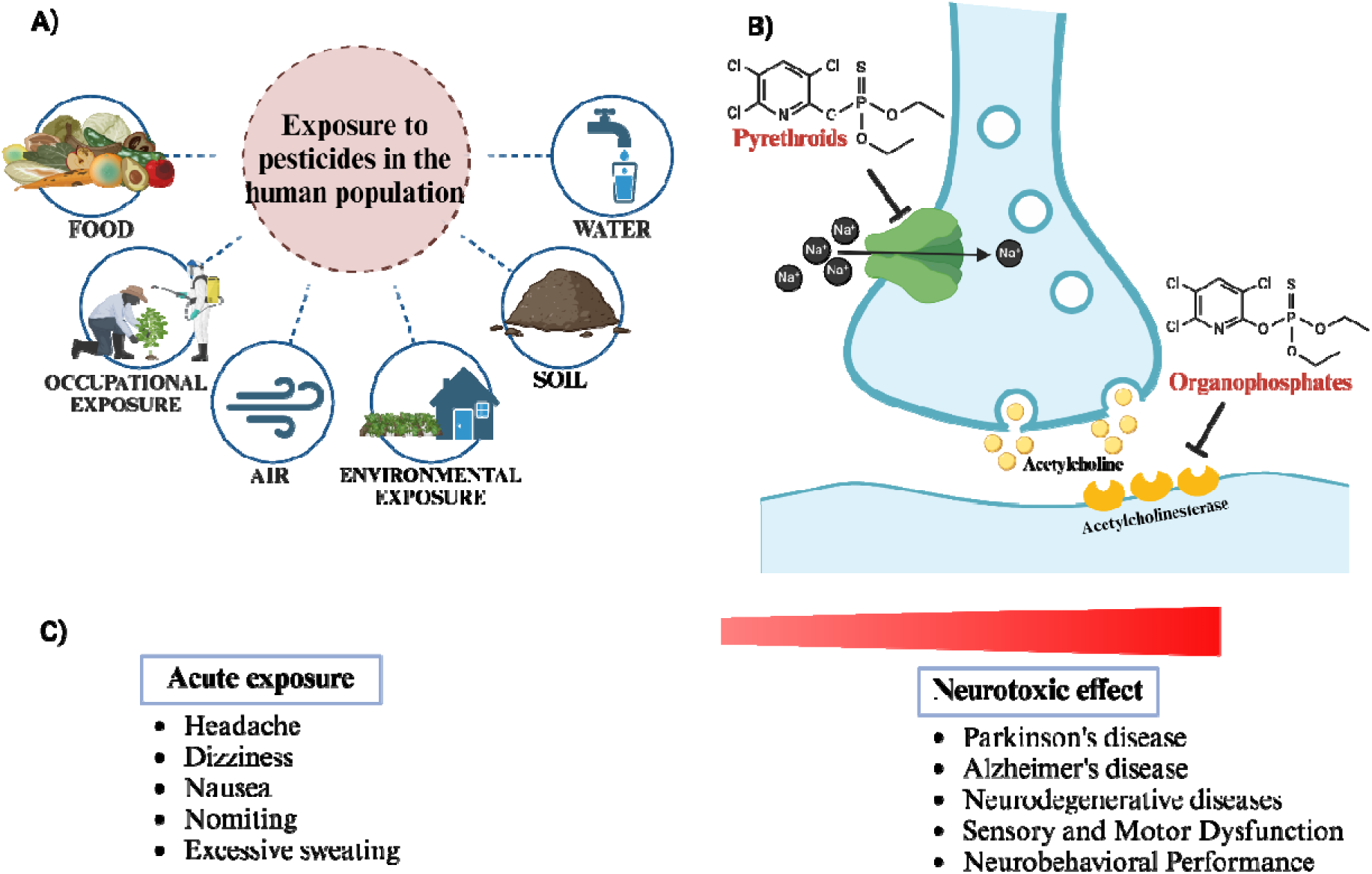
Exposure pathways and underlying neurological mechanisms associated with organophosphates (OP) and pyrethroids (PYR) pesticides A) Pesticide exposure pathways: OP and PYR can enter the human body through multiple routes, including inhalation, ingestion, dermal absorption, and contact with contaminated air, water, soil, and food. Agricultural workers are at higher risk due to occupational exposure, while the general population is primarily affected through environmental exposure. B) Molecular mechanisms of action: OP inhibit acetylcholinesterase at neuronal synapses, leading to the accumulation of acetylcholine and the overstimulation of postsynaptic cholinergic receptors. PYR disrupt neuronal function by altering sodium channel dynamics in the neuronal membrane, prolonging their opening and inducing abnormal neuronal excitability. C) Acute exposure to OP and PYR can result in symptoms such as headache, nausea, vomiting, excessive sweating, and tearing. Chronic exposure, however, is associated with long-term neurological consequences, including cognitive deficits, motor impairments, and an elevated risk of neurodegenerative diseases such as Parkinson’s and Alzheimer’s.

Pyrethroids, though less acutely toxic than OP, disrupt nerve function by altering sodium ion channel permeability in nerve membranes, which can lead to symptoms such as tingling, pruritus, and, in severe cases, tremors and seizures (Ahmad et al., 2024; Mohammadi et al., 2019).

Health risks associated with pesticide exposure are influenced by various factors, including application methods, frequency of use, protective measures, and the vulnerability of the exposed population (Tudi et al., 2022). Agricultural workers are particularly at risk due to their direct and prolonged exposure to these chemicals (Mostafalou and Abdollahi, 2013; Suratman et al, 2015). In developing countries, the lack of stringent regulatory oversight and limited awareness of pesticide safety practices further exacerbate exposure levels, compounding the associated health risks (Kubiak-Hardiman et al., 2023).

OP are well-documented in the literature for their acute neurotoxic effects on the cholinergic system, contributing to neurobehavioral difficulties and cognitive impairments (Muñoz-Quezada et al., 2013; Tsai et al., 2021). PYR, while considered less toxic than OP, have also been associated with an increased risk of neurological disorders due to their disruptive action on sodium channels (Antonangeli et al., 2023; Barr et al., 2010). While studies examining individual pesticide exposures provide robust evidence of their neurotoxicity (Lucero and Muñoz-Quezada, 2021), the combined effects of OP and PYR remain poorly understood, despite their frequent co-occurrence in both agricultural and residential environments (Bennette et al., 2020).

Additionally, growing concern focuses on the chronic effects of low-dose pesticide exposure on the general population, particularly its potential links to developmental disorders and neurodegenerative diseases such as Parkinson’s and Alzheimer’s (Baltazar et al., 2014). The risks to children are especially alarming, as their developing bodies and brains are significantly more vulnerable to the toxic effects of these chemicals (Rauh et al., 2012).

The widespread use of pesticides and the potential for broad exposure highlight the need for thorough evaluations of their health effects (Tang et al., 2021). Growing epidemiological evidence links pesticide exposure to various adverse outcomes, emphasizing the importance of rigorous research, stronger regulations, and international collaboration to support public health measures and promote safer alternatives in pest management

Several systematic reviews have analyzed the relationship between pesticides and brain health, primarily focusing on exposure to OP or PYR separately. Biosca-Brull et al. (2021) evaluated the association between different classes of pesticides and autism spectrum disorder (ASD), finding consistent links with OP but not examining their combination with PYR. Ongono et al. (2020) reviewed the effects of various pesticides on neurodevelopment, including OP and PYR, but did not address simultaneous exposure.

Burns et al. (2013) compiled studies on pesticides and neurodevelopment, mentioning some cases where both compounds coincided, but without analyzing them as a combined exposure. Other reviews have studied their relationship with neurodegenerative and psychiatric disorders (Soleman et al., 2024) without distinguishing the potential effects of their interaction in humans.

This systematic review updates the evidence on OP and PYR co-exposure and its relationship with neurobehavioral, psychiatric, and neurodegenerative disorders. Unlike previous studies, it focuses exclusively on epidemiological research that has assessed both exposures simultaneously, allowing for the identification of association patterns and research gaps.

This review addresses the following research question: What is the scientific evidence for the association between simultaneous exposure to OP and PYR pesticides and the prevalence of neurobehavioral difficulties, neurological disorders, and neurodegenerative diseases in children and adults?

This review aims "to analyze the scientific evidence from the past 10 years regarding simultaneous exposure to OP and PYR pesticides and their association with neurobehavioral difficulties, as well as neurodegenerative, and neurological disorders in humans". The global reliance on chemical pest control has led to an increased risk of combined exposure to multiple pesticides, presenting a significant public health challenge. To date, no systematic review has thoroughly examined the effects of simultaneous OP and PYR exposure, making this analysis both timely and scientifically significant. By synthesizing recent findings, this review seeks to provide a deeper understanding of the potential synergistic effects of these compounds on brain health. This evaluation endeavors to address a significant gap in toxicological research, contribute to the development of enhanced public health guidelines, and inform regulatory policies.

## Methods

To identify the most relevant scientific literature, we conducted a systematic review following the PRISMA (Preferred Reporting Items for Systematic Reviews and Meta-Analyses) guidelines. This rigorous approach ensures transparency and reproducibility in the selection process. The detailed PRISMA workflow, including study identification, screening, eligibility, and inclusion, is presented in Figure 2 (Page et al., 2021).

**Figure 2.**
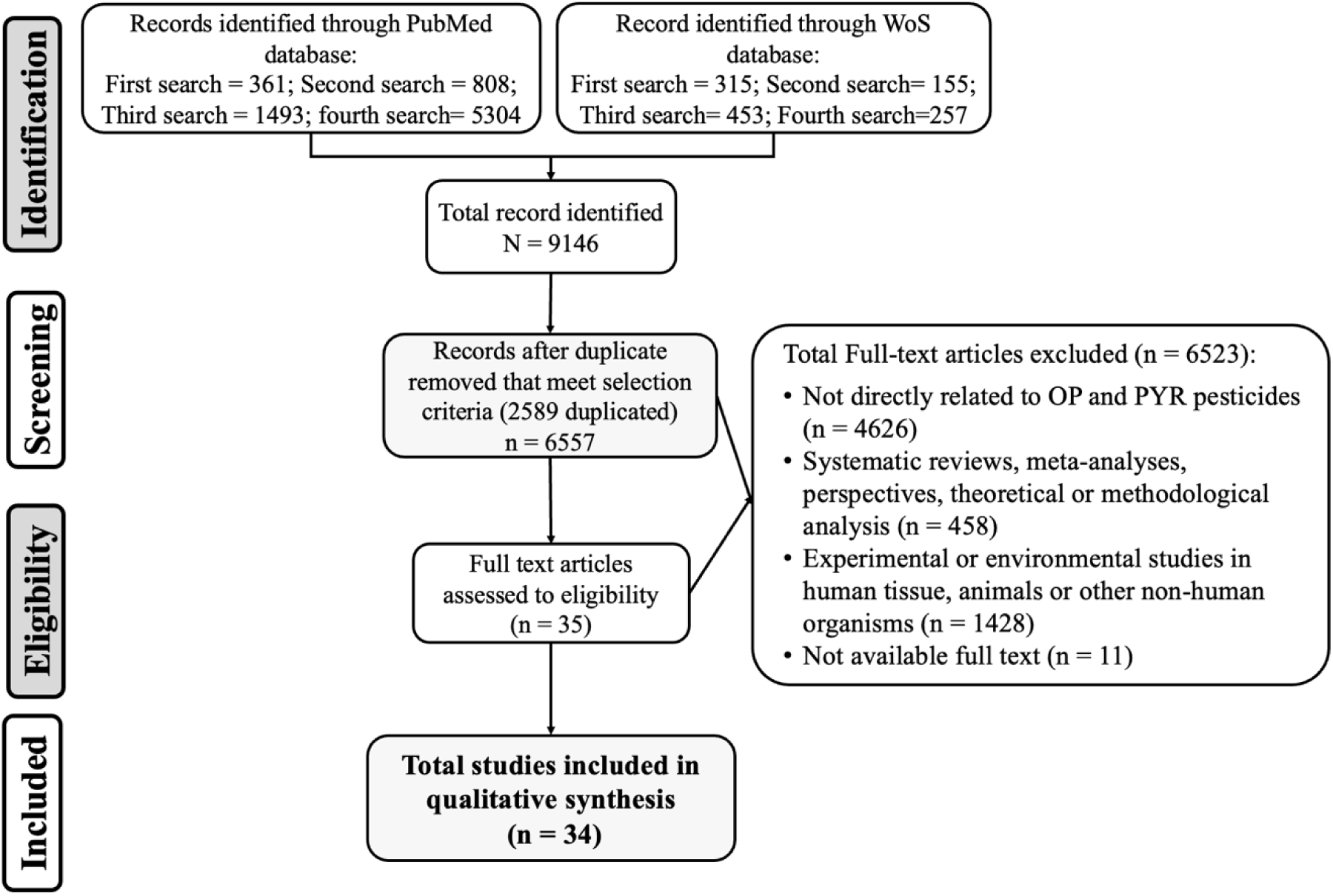
PRISMA flow diagram summarizing the identification, screening, eligibility, and inclusion of studies for the systematic review

The literature search was confined to the PubMed and Web of Science (WoS) databases, chosen for their extensive coverage in the field of epidemiology. Articles were limited to those published in English to ensure consistency and precision in data analysis.

Four comprehensive searches were conducted using the ’all fields’ option in both databases to capture a broad range of relevant studies. The initial search used the terms (pesticides) AND (neurobehavioral) to focus on primary research and exclude review articles. The second search employed the terms (pesticides) AND (mental disorders), followed by a third search with (pesticides) AND (neurodegenerative diseases). Finally, the fourth search utilized (pesticides) AND (neurological disorders) to identify primary research articles examining the relationship between OP and PYR pesticide exposure and brain health.

Review articles were systematically excluded across all searches to ensure a focus on original research findings. Studies published between 2014 and 2024 were prioritized to capture the most recent and relevant evidence. The databases were last consulted on December 1, 2024, ensuring the inclusion of up-to-date studies.

This review focuses on epidemiological studies investigating the relationship between simultaneous exposure to OP and PYR pesticides and neurobehavioral performance, mental health disorders, neurodegenerative diseases, and other neurological conditions in children and adults. Studies that did not explicitly examine the interaction between OP and PYR pesticides and the risk of neurological or related disorders were excluded. Additionally, literature reviews, commentaries, editorials, and animal-based observational or environmental studies were omitted, as they fall outside the scope of this analysis.

A team of five researchers conducted the search and selection of manuscripts collaboratively. Following a systematic procedure aligned with the predefined criteria, the team worked to identify and evaluate relevant studies. Regular meetings were held to compare findings and ensure unanimous agreement on the inclusion of each study, achieving a 100% consensus before proceeding. Subsequently, 13 of the authors validated the review. The selected studies were then systematically organized based on the evidence they provided regarding the relationship between pesticide exposure and brain health.

Epidemiological studies were evaluated using the ROBINS-E tool (Risk of Bias in Non-Randomized Studies of Exposures), which provides a structured and systematic method to assessing study quality in observational cohort studies (Higgins et al., 2024).

The evaluation process considered seven bias domains: confounding, exposure measurement, participant selection, post-exposure interventions, missing data, outcome measurement, and selection of reported results. Each domain was analyzed to determine its potential impact on study validity. The classification of bias risk in each domain followed the tool’s criteria, categorizing studies as low, some concerns, high, or very high risk (Higgins et al., 2024).

Confounding (Domain 1) was assessed by identifying external factors such as socio- demographic characteristics, environmental influences, and occupational exposure, which could affect both exposure and outcome, potentially distorting the observed association. Exposure measurement (Domain 2) was evaluated in terms of accuracy and reliability, considering potential classification errors and recall bias. Participant selection (Domain 3) was examined to ensure comparability between exposed and unexposed groups, preventing systematic selection biases.

For post-exposure interventions (Domain 4), the analysis considered whether participants, upon becoming aware of their exposure, modified their behavior or accessed medical treatments, potentially influencing outcomes and introducing bias if these changes were unevenly distributed across groups (Table 1). Missing data (Domain 5) was examined concerning participant attrition or incomplete records, assessing their impact on the validity of findings. Outcome measurement (Domain 6) was evaluated in terms of consistency and accuracy in assessing neurological health effects, ensuring the uniform application of diagnostic criteria. Lastly, selection bias in reported results (Domain 7) was analyzed to determine whether certain findings were prioritized over others, which could influence the overall interpretation of results.

**Table 1.**
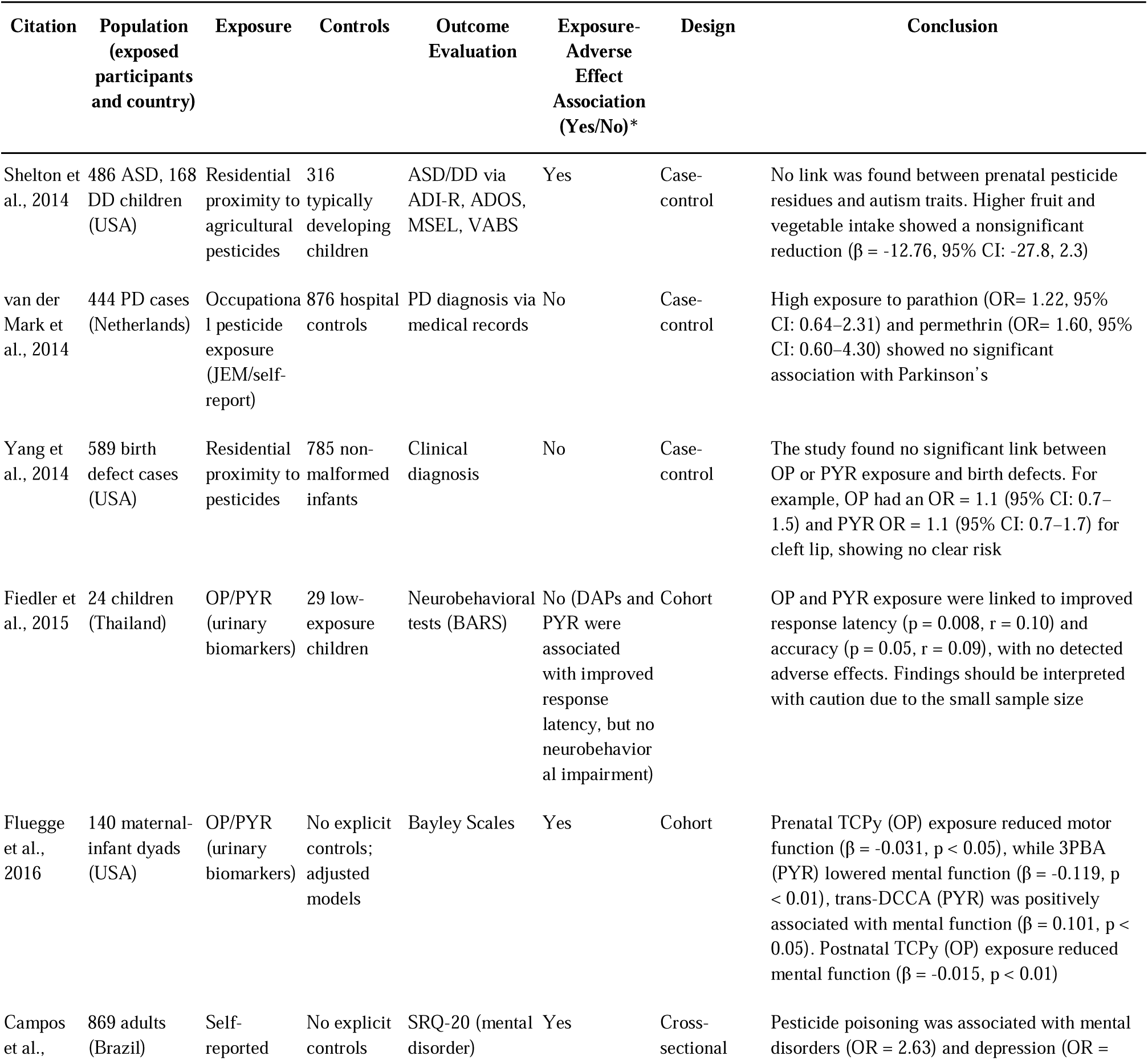
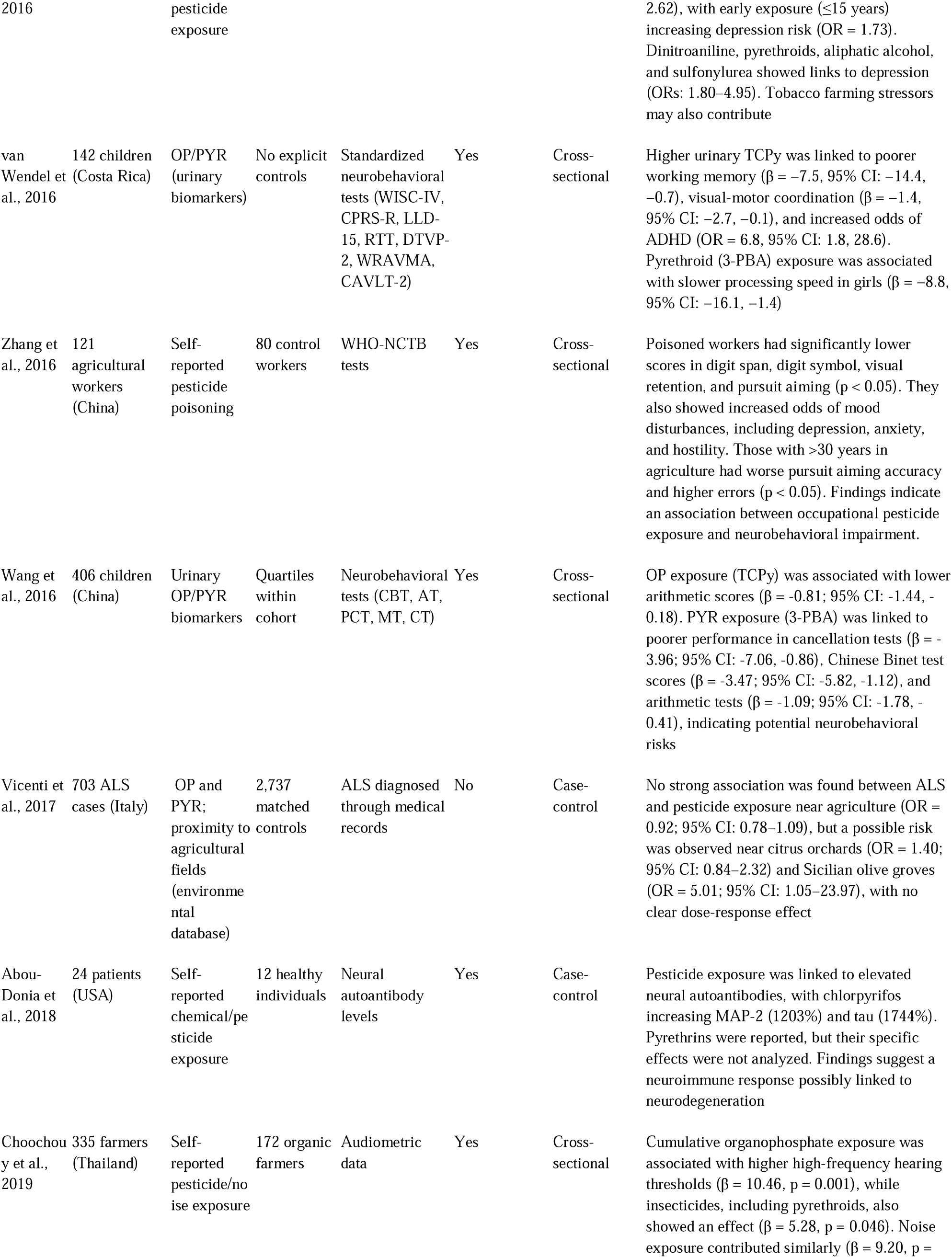
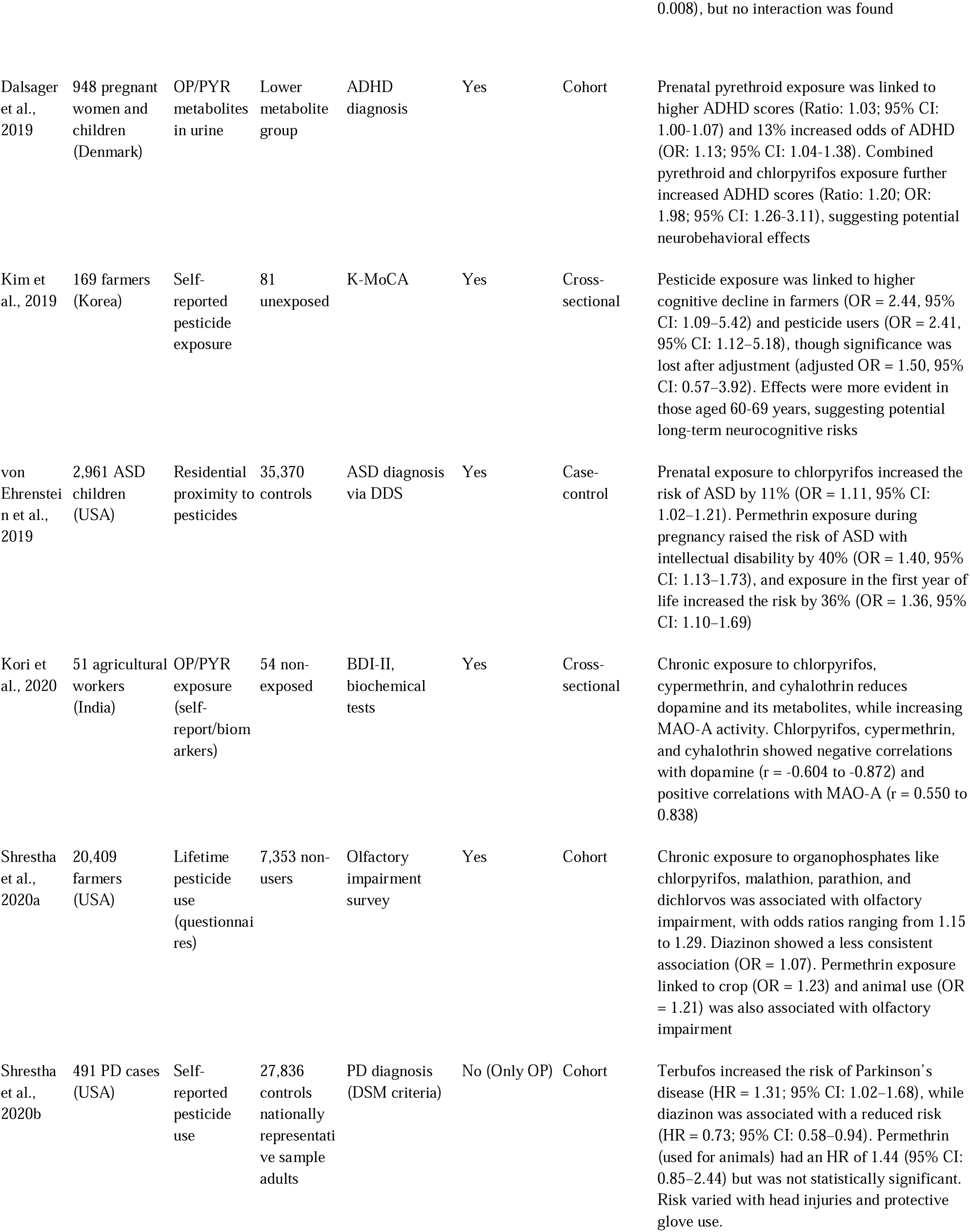
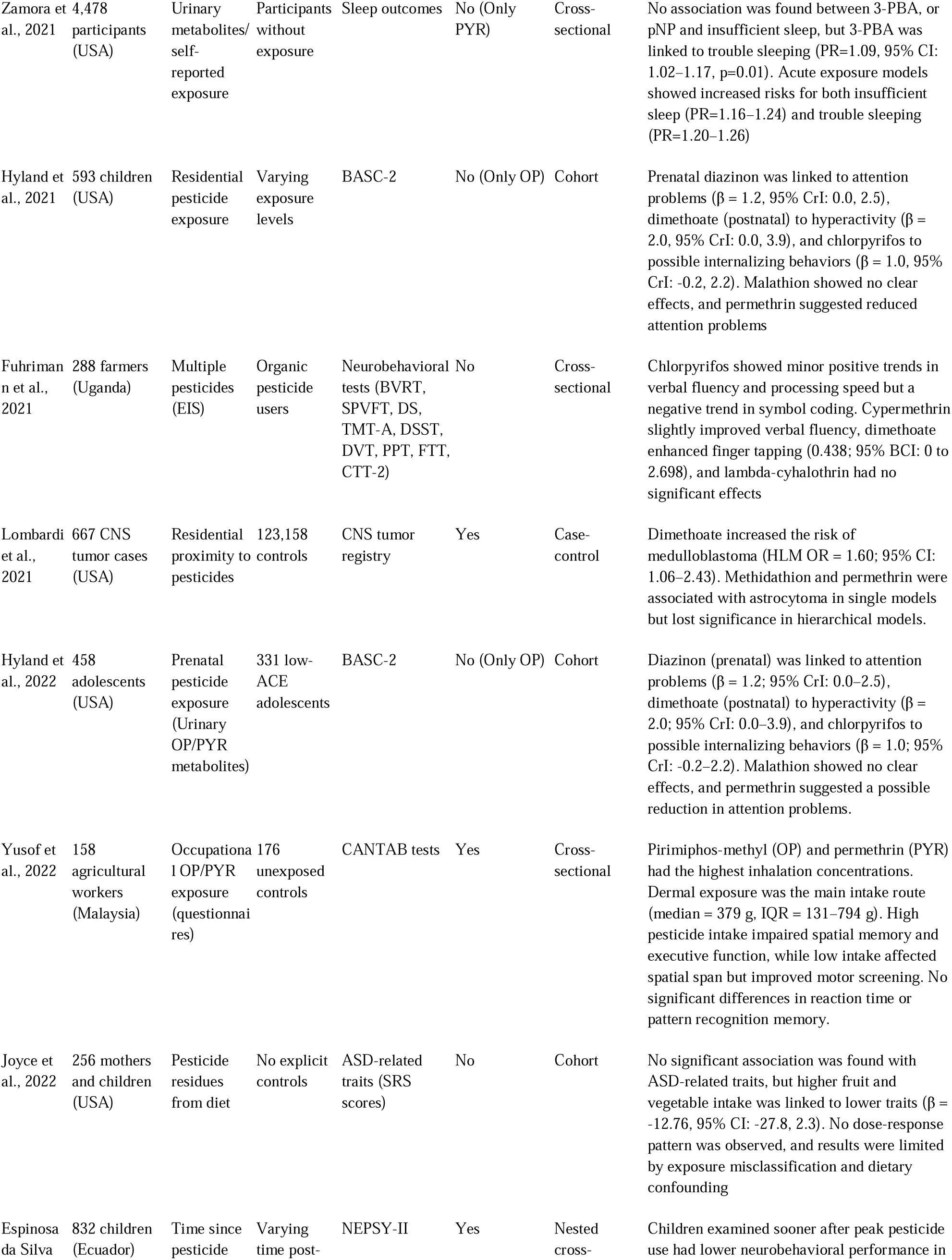
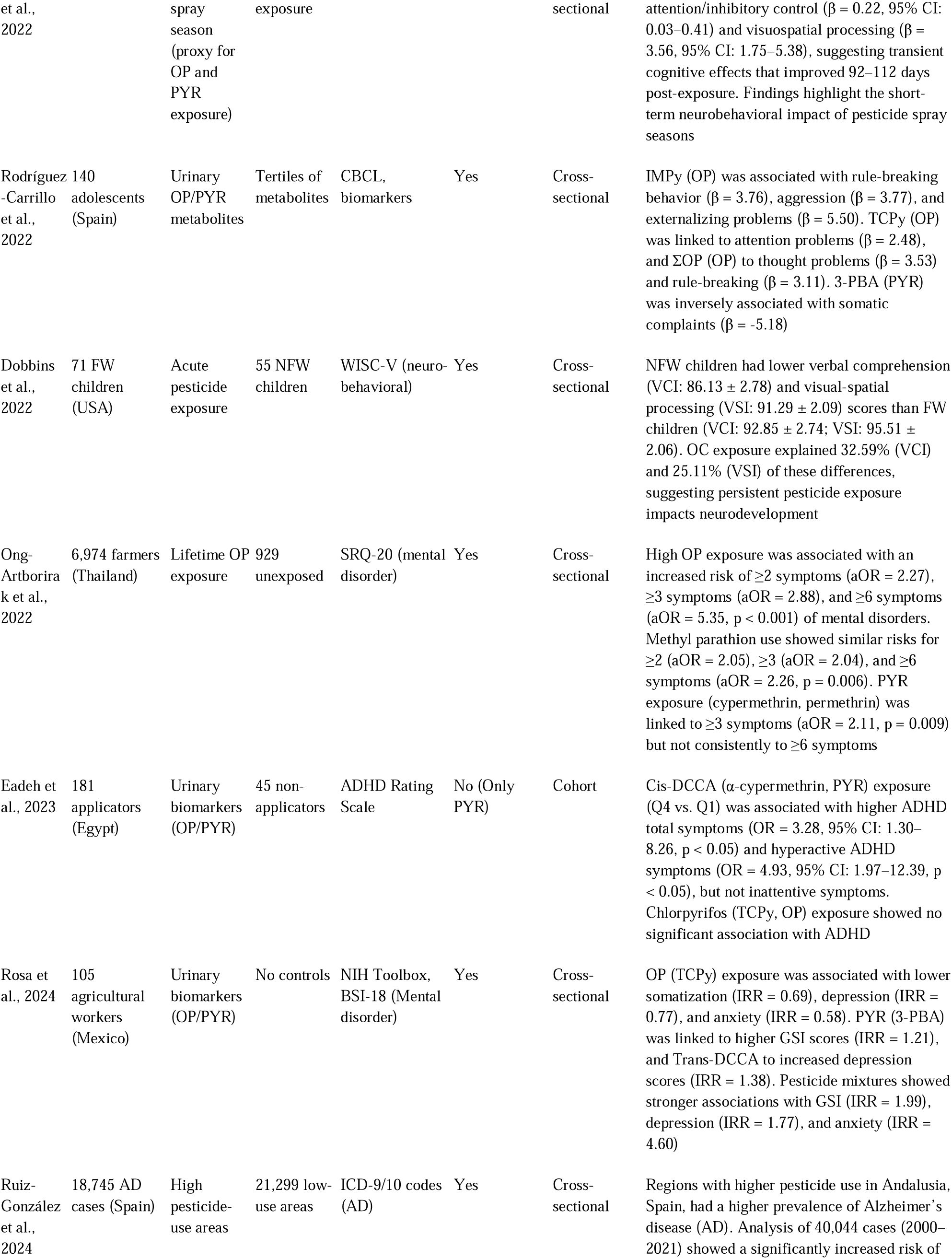
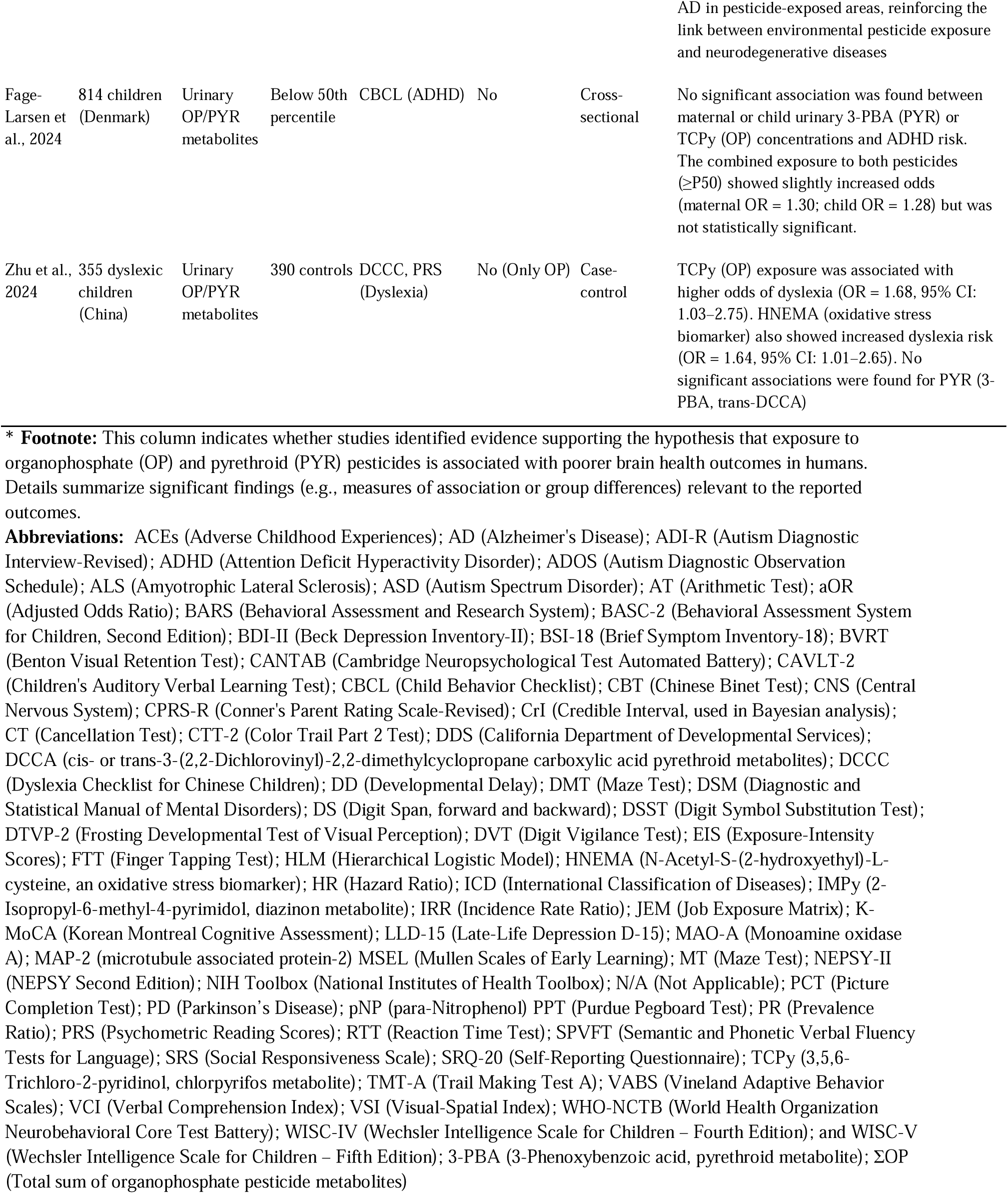
Summary of studies on simultaneous exposure to organophosphate (OP) and pyrethroid (PYR) pesticides, categorized by the PECO framework and assessed for neurobehavioral performance, neurological disorders, neurodegenerative diseases, and mental health outcomes using the ROBINS-E tool for cohort studies

Beyond bias assessment, the PECO framework (Population; Exposure; Comparator; and Outcomes) (Morgan et al., 2018) was applied for the systematic synthesis of included studies. This model facilitated the categorization of evidence by considering population, exposure, control groups, and measured outcomes, enabling an evaluation of methodological quality and the strength of reported findings. The characteristics of each study, including design, sample size, exposure methods, and outcome assessment, are detailed in Table 1.

Table 2 presents statistical measures from nine cohort studies evaluating the association between simultaneous OP and PYR exposure and brain health outcomes. It includes effect estimates such as odds ratios, hazard ratios, and Beta coefficients, with confidence intervals to differentiate significant from non-significant findings. In contrast, Table 3 provides a categorical summary of these studies by brain health outcome, indicating the presence or absence of associations without reporting specific statistical values.

**Table 2.**
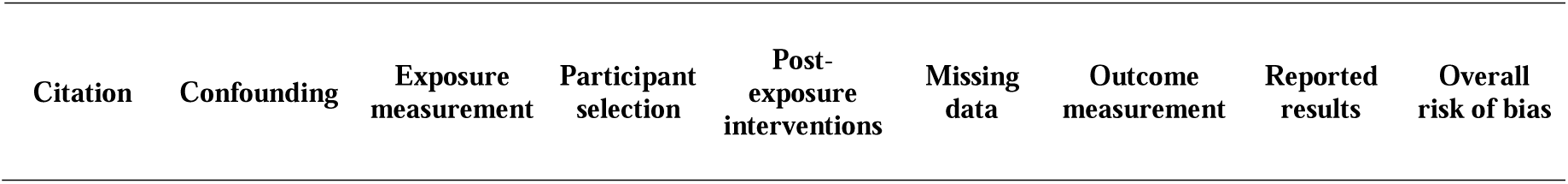
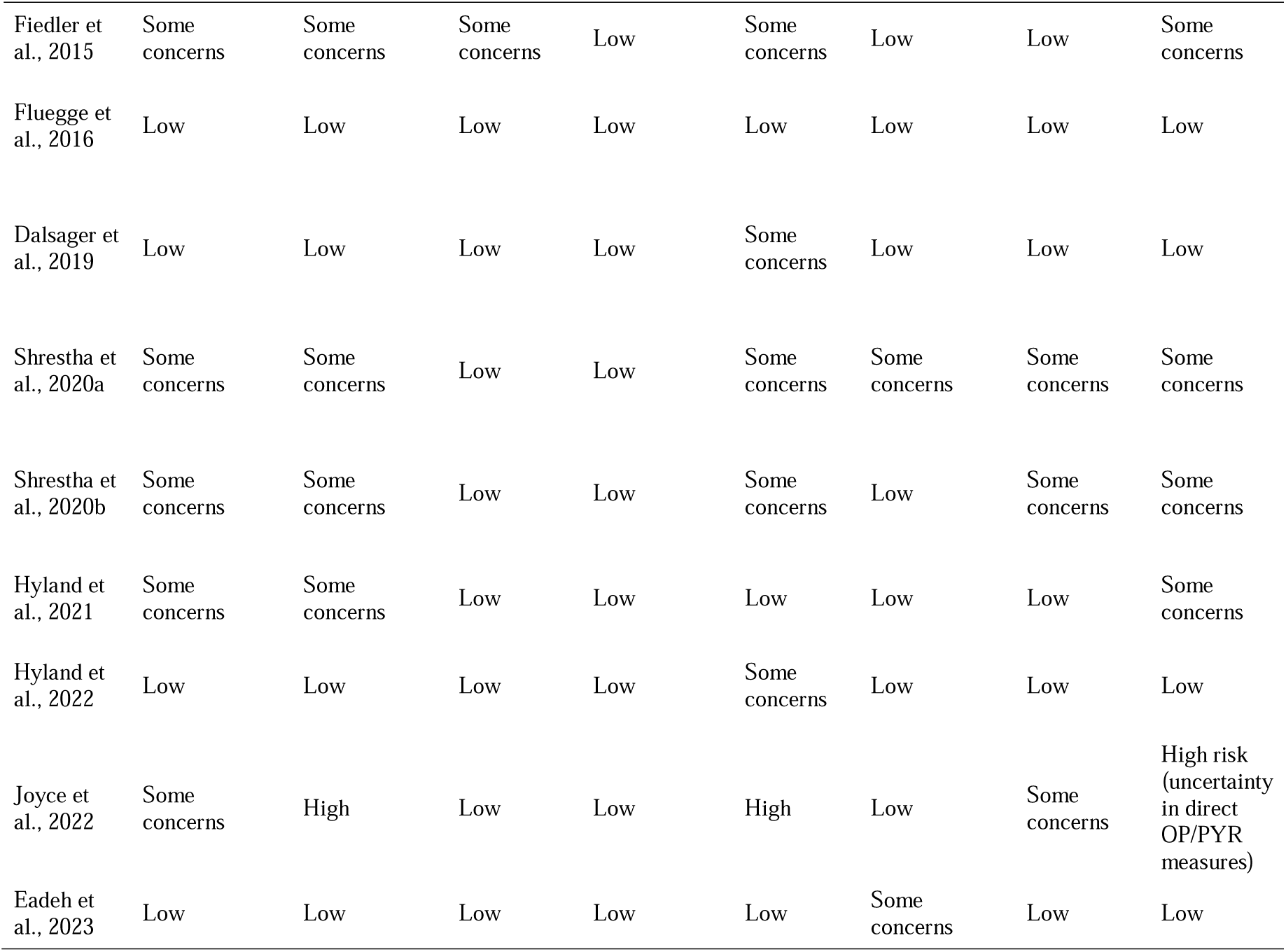
Risk of bias assessment across the seven ROBINS-E domains and overall risk in cohort studies on OP and PYR exposure.

**Table 3.**
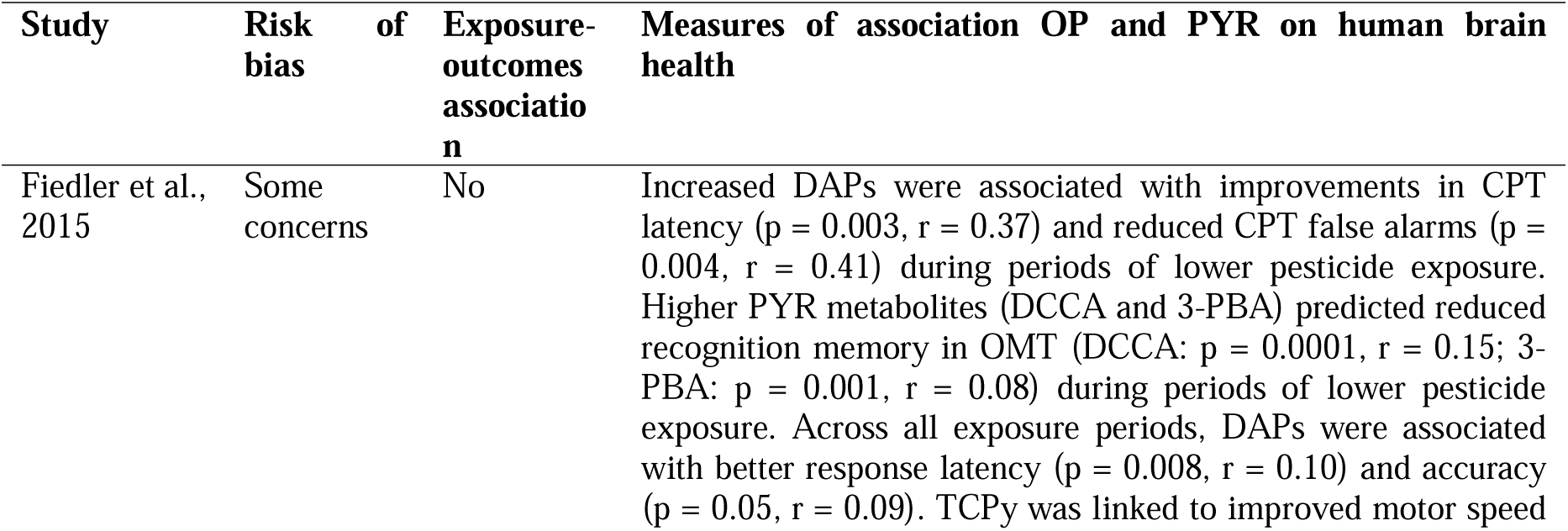
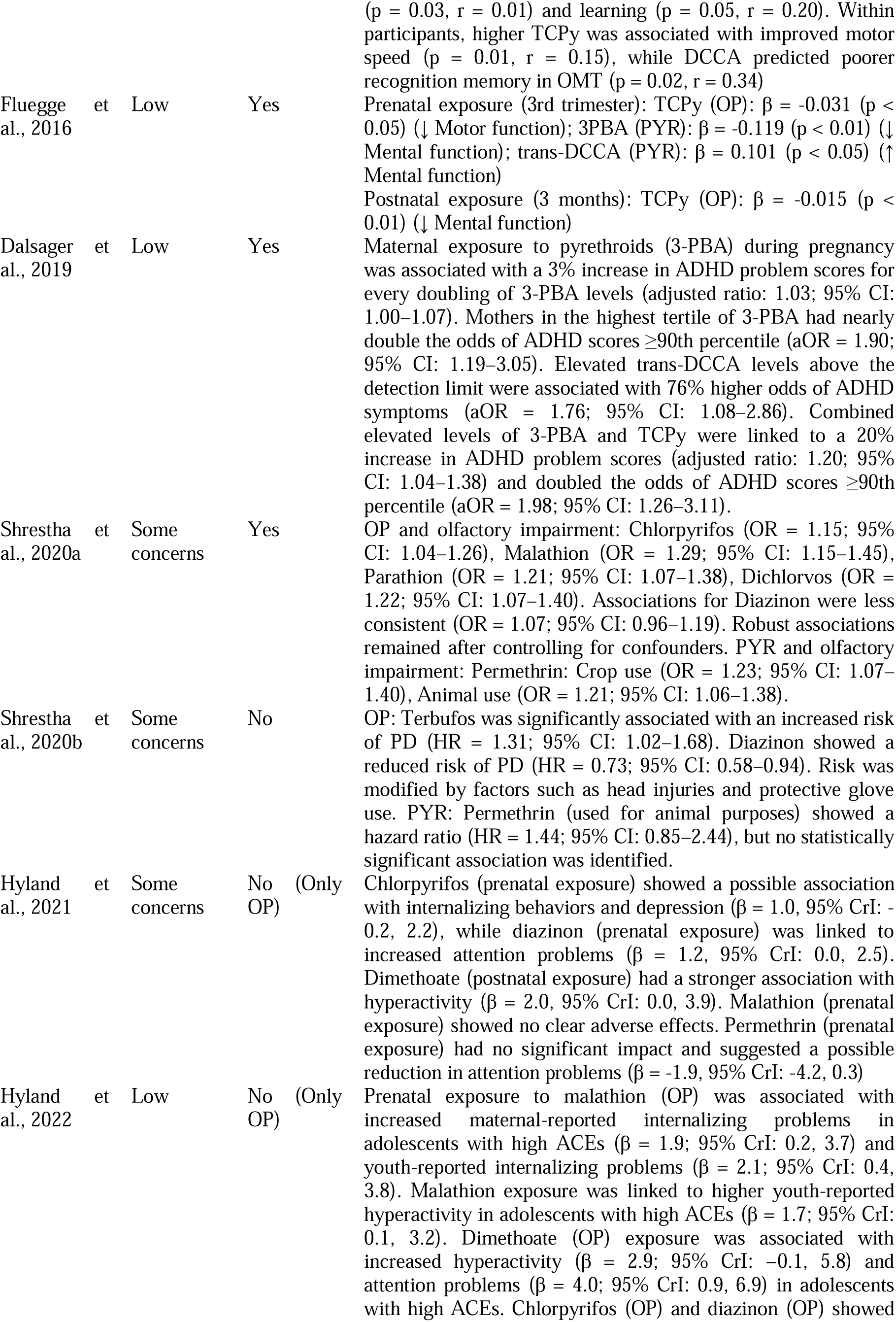
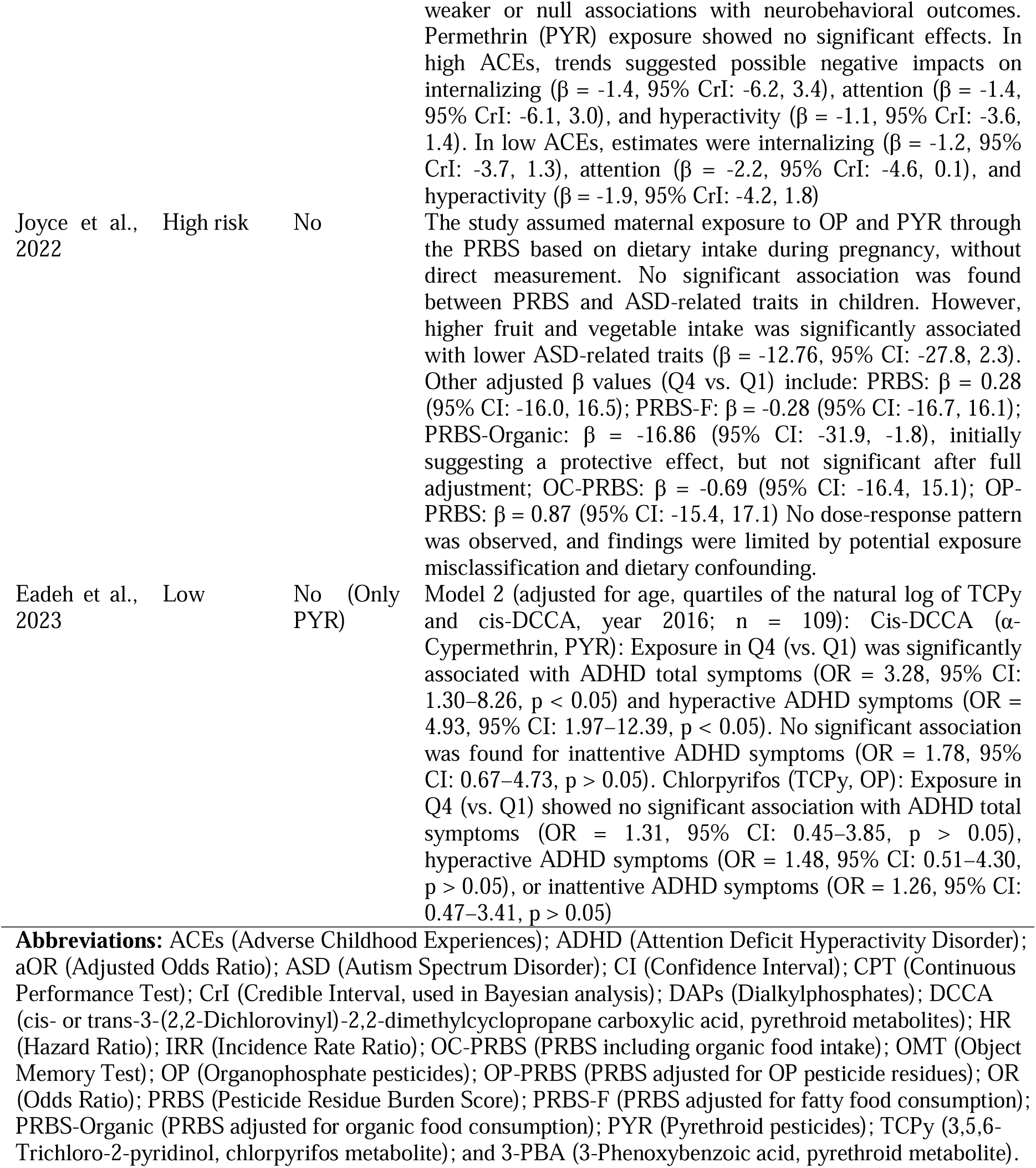
Association measures from cohort studies (n = 9) on the effects of simultaneous organophosphate (OP) and pyrethroid (PYR) exposure on brain health outcomes.

Figures 3 and 4 from cohort studies, using Beta coefficients and odds ratios (ORs), respectively, to assess the association between combined OP and PYR exposure and neurological health outcomes. Cohort studies were prioritized, but methodological heterogeneity in statistical analyses precluded a meta-analysis.

**Figure 3.**
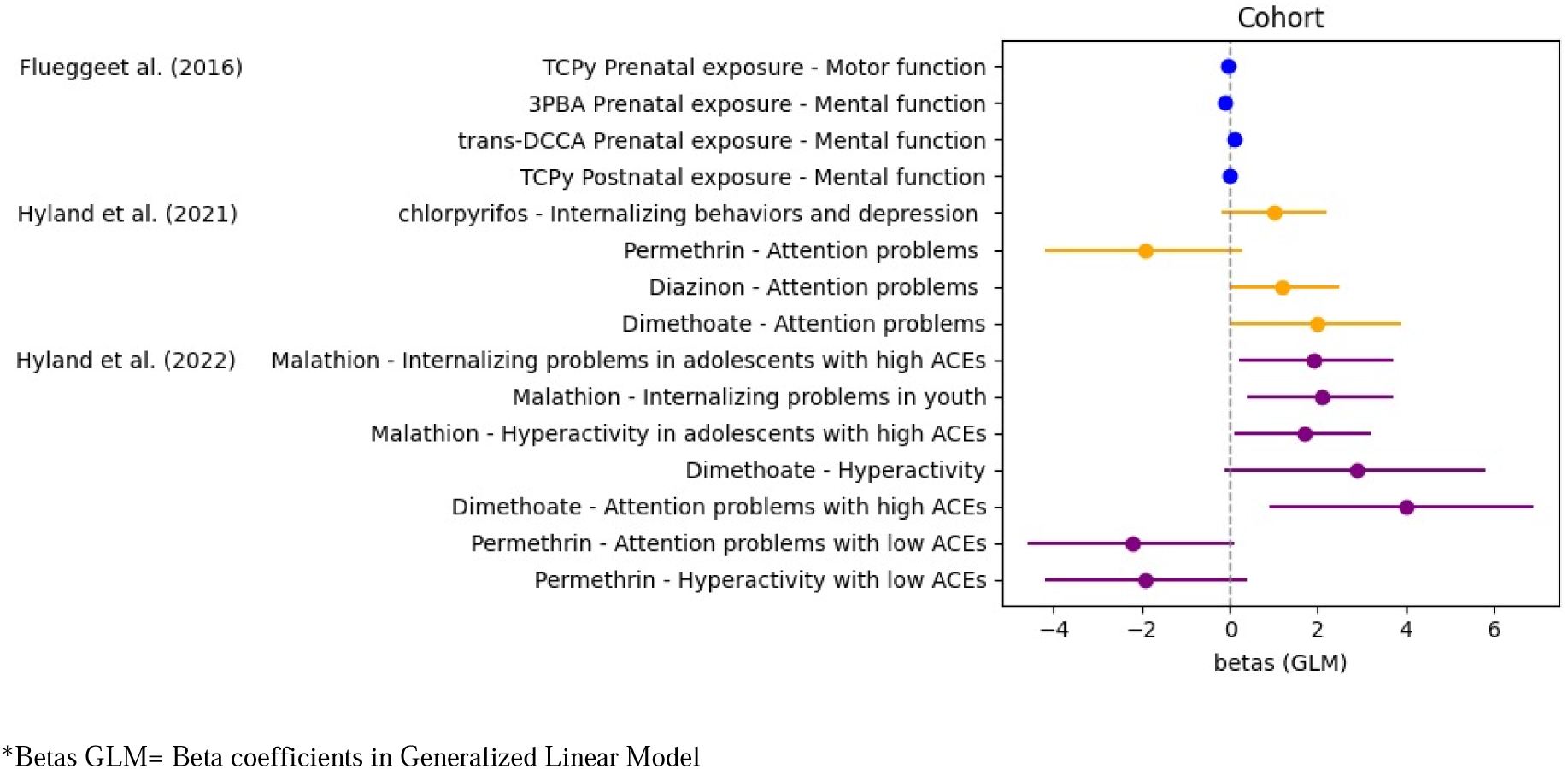
Association between organophosphate (OP) and pyrethroid (PYR) pesticide exposure and brain health outcomes based on the three reviewed studies (Betas GLM, CI 95%)*

**Figure 4.**
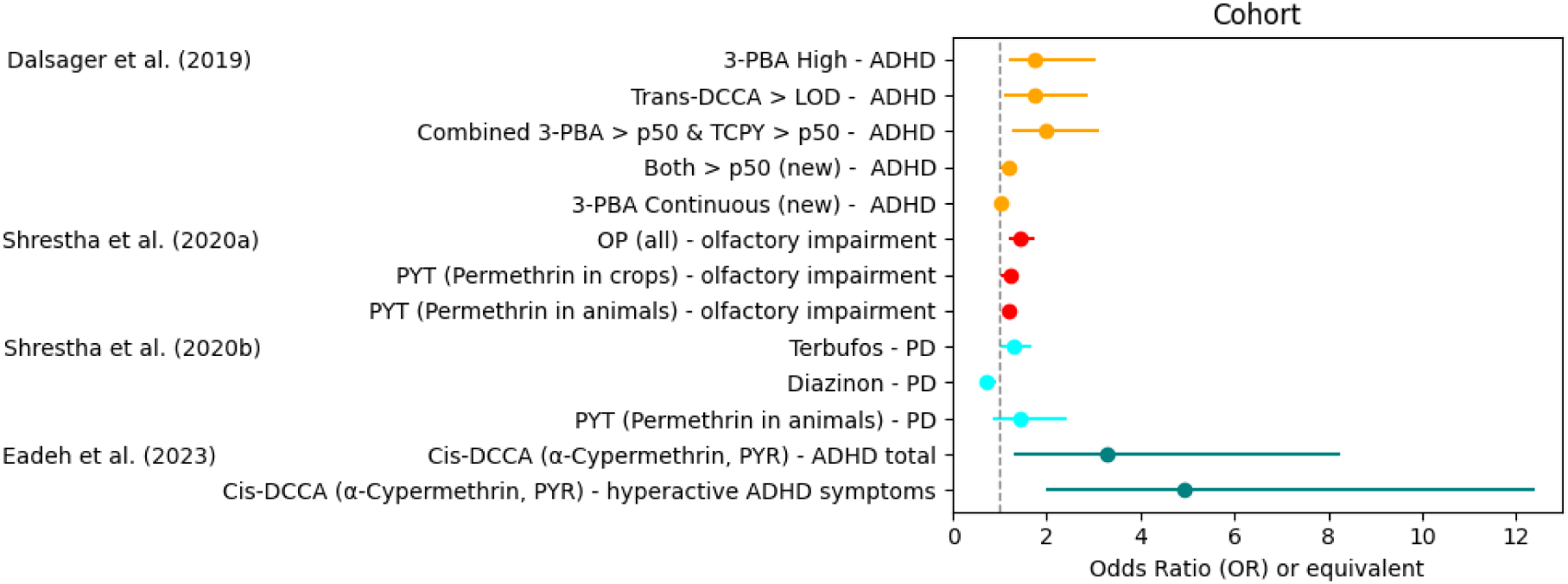
Association between pesticide exposure and brain health outcomes using Odds Ratios (OR, CI 95%)

## Results

The included studies are summarized in Figure 1 and Table 1, which categorize them according to the PECO framework (Morgan et al., 2018) for all studies and the ROBINS-E (Higgins et al., 2024) evaluation criteria for cohort studies only.

Of the 34 studies included in this systematic review (Table 1), the majority were conducted in North America (38%, 13 studies), followed by Europe (18%, 6 studies), Asia (26%, 9 studies), Latin America (12%, 4 studies), and Africa (6%, 2 studies). This distribution indicates a higher concentration of research in regions with advanced capabilities for assessing pesticide exposure and its effects on human health, or where researchers have established collaborations with well-equipped institutions to conduct such analyses.

Regarding the populations analyzed, 41% of the studies (14 out of 34) focused on children, who are particularly vulnerable to neurotoxic effects due to their developing nervous systems. The remaining 53% (18 out of 34) examined adults, with a significant focus on agricultural workers experiencing chronic exposure. Additionally, 73% of the studies (n = 25) included sample sizes exceeding 200 participants, enhancing the statistical power of their findings, while 27% utilized smaller sample sizes.

Methods for measuring pesticide exposure varied across the studies, 41% (n = 14) utilized specific biomarkers in blood and urine, considered the gold standard for accuracy, while 32% relied on questionnaires and self-reports, which, though practical, carry a higher risk of information bias.

For assessing the effects of exposure, 14 studies (41.2%) employed neurobehavioral scales, 10 (29.4%) utilized checklists, questionnaires, or surveys, 8 (23.5%) relied on clinical diagnoses, and 2 (5.9%) used biomarkers for outcome evaluation.

The most common study design was the analytical cross-sectional approach, utilized in 50% (17 out of 34) of the studies, due to its practicality in assessing exposure and effects simultaneously. Cohort studies accounted for 26% (9 out of 34), providing a more robust analysis of changes over time, while case-control designs made up 24% (8 out of 34).

Across the 34 studies examining simultaneous exposure to OP and PYR pesticides, 62% (21 studies) reported an association with adverse brain health effects. Additionally, 6% (2 studies) found no association with OP but did with PYR, while 12% (4 studies) found an association with OP but not with PYR. Of these 21 studies that found an association with adverse brain health effects, 14% were cohort studies (3 studies), 19% were case-control studies (4 studies), 62% were cross-sectional studies (13 studies), and 5% corresponded to a nested cross-sectional study (1 study).

Regarding the risk of bias, among the nine cohort studies, one was classified as high risk, primarily due to difficulties in directly assessing exposure to OP and PYR, which made it harder to establish a dose-response relationship. Additionally, this study lacked a clearly defined control group. Four studies were categorized as having some concerns, either due to a limited sample size or the reliance on indirect methods for exposure assessment instead of biomarker-based evaluations.

Four studies were considered to have a low risk of bias. Among the cohort studies, only three reported an association between simultaneous exposure to OP and PYR, with one classified as having some concerns and two as low risk. Among the studies that examined only OP, three identified an association, two of which had some concerns.

Of the two cohort studies that did not find an adverse association between OP and PYR exposure and brain health, one was classified as high risk due to the lack of direct pesticide exposure assessment, while the other was categorized as having some concerns, mainly due to sample size limitations. Table 2 summarizes the risk of bias assessment for the nine cohort studies using the ROBINS-E tool.

Table 3 summarizes the association measures from the nine cohort studies, detailing their findings on the relationship between OP and PYR exposure and adverse brain health outcomes.

Of the 9 studies analyzed, 3 identified an association between simultaneous exposure to OP and PYR pesticides and adverse effects on human brain health. These findings are summarized in the categories presented in Table 4.

**Table 4.**
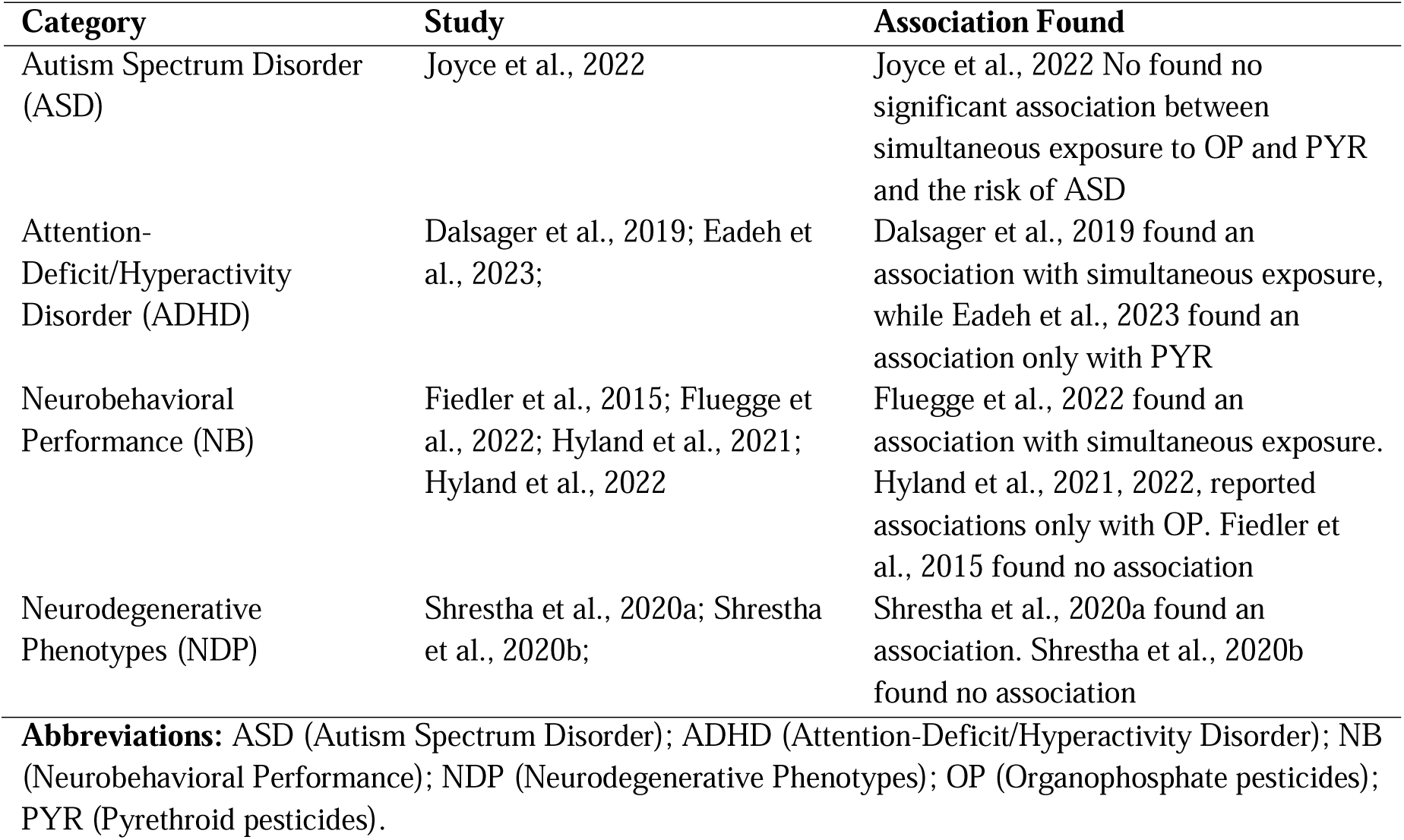
Summary of associations between simultaneous exposure to organophosphate (OP) and pyrethroid (PYR) pesticides and brain health outcomes (9 studies)

The table 4 summarizes findings from various studies examining the association between pesticide exposure, specifically OP and pyrethroids PYR, and neurological conditions. In the case of ASD, Joyce et al. (2022) reported no association between simultaneous exposure to OP and PYR and an increased risk of ASD.

Regarding ADHD, the findings were mixed. Dalsager et al. (2019) identified an association with simultaneous exposure to OP and PYR, whereas Eadeh et al. (2023) found an association only with PYR suggesting that pyrethroids may have a impact on ADHD risk. For NB, studies have shown mixed results depending on the type of pesticide exposure.

Fluegge et al. (2022) reported a link between simultaneous OP and PYR exposure and neurobehavioral impairments. In contrast, Hyland et al. (2021, 2022) found associations only with OP, suggesting that OP may have a stronger impact on neurobehavioral outcomes. Fiedler et al. (2015) did not find adverse effects from OP and PYR exposure; instead, they observed a positive association.

In the case of NDP, results were inconsistent. Shrestha et al. (2020a) found a significant association between pesticide exposure and neurodegenerative outcomes, while Shrestha et al. (2020b) did not observe any relationship, highlighting the need for further investigation. Overall, the findings suggest potential neurotoxic effects of OP and PYR exposure, particularly in ADHD and neurobehavioral impairments. However, differences in exposure type and study design may contribute to the variability in reported associations, especially in the context of neurodegenerative diseases. Further research is necessary to clarify these relationships and better understand the mechanisms underlying pesticide-related neurological effects.

Although a meta-analysis was not performed due to methodological heterogeneity among the studies.

Figure 3 presents the measures of association (Beta coefficients) and 95% confidence intervals (CI95%) between pesticide exposure and brain health outcomes in cohort studies with moderate or low risk of bias. The forest plot depicts these associations based on the studies by Fluegge et al. (2016) and Hyland et al. (2021, 2022).

Figure 4 displays the Odds Ratios (OR, CI95%) for the association between pesticide exposure and brain health outcomes in cohort studies with moderate or low risk of bias. The forest plot illustrates findings from Dalsager et al. (2019), Shrestha et al. (2020a, 2020b), and Eadeh et al. (2023), providing additional insight into the potential risks associated with pesticide exposure.

Figures 3 and 4 show the variability in findings across different brain health outcomes, reflecting differences in study design, outcome measures, and exposure assessments. This heterogeneity emphasizes the need for further research to refine these associations and address existing gaps in the evidence.

## Discussion

This review examines studies from the last ten years that have investigated simultaneous exposure to OP and PYR pesticides and their impact on brain health, including cohort, case-control, and cross-sectional designs. However, when assessing methodological quality, it is recognized that the ROBINS-E tool is particularly effective for cohort studies, as it allows for a more detailed evaluation of bias in longitudinal observational research. In contrast, its application to case-control and cross-sectional studies has limitations, which may affect the interpretation of findings and comparability across study designs (Higgins et al., 2024).

By leveraging this tool, we identified and analyzed potential biases related to confounding factors, selection criteria, and exposure measurements, enhancing the reliability of the findings.

The analysis suggests a potential association between combined OP and PYR pesticide exposure and brain health outcomes, including neurobehavioral disorders, neurodegenerative diseases, and psychiatric conditions. However, findings varied across cohort studies, depending on the risk of bias, exposure assessment methods, and study design. Some studies, such as Fluegge et al. (2016) and Dalsager et al. (2019), which had low risk of bias and biomarker-based exposure assessments, reported significant associations, particularly with neurodevelopmental and attention-related outcomes. In contrast, some studies despite having a large sample size, relied on self-reported exposure data, which may produce memory bias and affect the strength of associations. In adults, cohort studies examining chronic occupational or residential pesticide exposure provided evidence of neurodegenerative risks, but findings were inconsistent (Shrestha et al., 2020a, 2020b). These inconsistencies may be due to variations in exposure assessment methods, as biomarker-based studies tend to produce stronger associations than self-reported exposure data.

In the case-control studies presented in Table 1, Vicenti et al. (2017) and van der Mark et al. did not find an association between simultaneous pesticide exposure and neurodegenerative diseases, specifically ALS (Vicenti et al., 2017) and PD (van der Mark et al., 2014). In contrast, Lombardi et al. (2021) reported a link between residential pesticide exposure and an increased risk of CNS tumors.

No cohort study included in this review identified an association between simultaneous OP and PYR exposure and mental health outcomes. However, evidence from other study designs suggests a potential link. For instance, Kori et al. (2020) reported a correlation between OP exposure and depressive symptoms, while Ong-Artborirak et al. (2022) and Rosa et al. (2024) observed associations between pesticide exposure and disruptions in neurotransmitter pathways. These findings suggest that cohort studies may not have adequately captured the mental health effects, possibly due to differences in study design, exposure assessment methods, or outcome definitions. Future research should incorporate standardized psychiatric assessments and biomarker-based exposure measures to better elucidate the relationship between pesticide exposure and mental health outcomes.

These findings indicate that combined pesticide exposure may have diverse effects, influenced by study design and exposure assessment methods. Considering psychosocial factors in future research could help clarify their role in neurodevelopmental and mental health outcomes, particularly in populations with sustained or high exposure levels.

Assessing the combined effects of OP and PYR pesticides accounts for real-world exposure, where multiple compounds often interact. Acetylcholinesterase inhibition by OPs like malathion and dimethoate may contribute to neurotoxic effects by disrupting neural signaling (Hyland et al., 2022). Considering pesticide mixtures in research improves understanding of their potential health impacts, particularly in populations with prolonged or high exposure.

Despite its contributions, this review has limitations. Variability in study designs, exposure metrics, and outcome measures complicates cross-study comparisons. While some studies used biomarkers for exposure assessment, others relied on self-reported exposure data, increasing the risk of recall bias and misclassification.

Furthermore, not all studies adequately adjusted for confounding factors such as socioeconomic status and environmental variables, which may have influenced the observed outcomes (Morgan et al., 2018). These limitations underscore the urgent need for standardized methodologies in future research.

Publication bias remains a concern, as studies reporting negative or null results may be underrepresented, potentially affecting the interpretation of associations (Higgins et al., 2024). To minimize this, we prioritized studies classified as high-quality based on ROBINS-E. While this approach improves the reliability of findings, ROBINS-E needs further refinement to be effectively applied in case-control and retrospective studies, where exposure assessment and bias control are more complex.

Finally, the geographical scope of the reviewed studies was limited, with most conducted in North America and Europe. Regions such as Africa and Southeast Asia, where pesticide use patterns and regulatory frameworks differ significantly, were underrepresented. Expanding research to these areas is critical for capturing variations in pesticide exposure and health outcomes on a global scale (Tudi et al., 2022).

For policymakers, this review emphasizes the need for stricter regulations on pesticide mixtures, as current frameworks often assess chemicals individually, overlooking the cumulative risks of co-exposure. Promoting integrated pest management strategies and supporting the development of safer pesticide alternatives are essential steps to mitigate these risks (Silva-Neto et al., 2024). Additionally, raising awareness among agricultural workers about the health risks of pesticide exposure and encouraging the consistent use of personal protective equipment are practical measures to reduce individual risk (Mostafalou and Abdollahi, 2013).

Further research should prioritize longitudinal studies to evaluate the long-term health effects of pesticide exposure, incorporating biomarkers for more precise exposure assessment. Expanding studies to regions with high pesticide use and weaker regulatory oversight is crucial for assessing the global burden of pesticide-related neurotoxicity. Standardizing outcome measures, particularly for neurobehavioral and psychiatric effects, will improve cross-study comparability and support more robust meta-analyses. Moreover, the neurotoxic mechanisms of combined pesticide exposure (such as oxidative stress, neuroinflammation, and epigenetic modifications) remain insufficiently studied.

Investigating these mechanisms in the context of concurrent pesticide exposure is vital for elucidating the biological pathways underlying the observed effects and informing targeted interventions (Tang, 2020).

This review provides evidence of associations between simultaneous exposure to organophosphate and pyrethroid pesticides and adverse brain health outcomes, particularly neurodevelopmental effects such as attention deficits. Cohort studies with biomarker-based assessments reported consistent links, while findings for neurodegenerative diseases were less conclusive. Variability in study designs and exposure assessment methods underscores the need for standardized methodologies and long-term research to better characterize these risks. These findings emphasize the need to strengthen public health strategies aimed at reducing pesticide exposure and mitigating potential neurotoxic effect.

## Data Availability

The data used in the systematic review are derived from publicly available sources (PubMed and Web of Science). Supplementary datasets could be available only from the corresponding author on reasonable request

https://osf.io/qjv6m/

## Author Contributions

Conceptualization, M.T.M.-Q.; methodology, M.T.M.-Q., R.H., J.T., J.N., B.L., L.Z., C.V., A.C, R.C., J.P.G., B.C., N.L., C.C., J.A., P.O., and T.B.; investigation, M.T.M.-Q., R.H., J.T., J.N., and B.L.; software: M.T.M.-Q., B.L., L.Z., B.C., and N.L.; visualization: M.T.M.-Q., B.L., L.Z., B.C., N.L., P.O., and T.B; validation: M.T.M.-Q., B.L., L.Z., C.V., A.C, R.C., J.P.G., B.C., N.L., C.C., J.A., P.O., and T.B; writing—original draft preparation, M.T.M.-Q., R.H., J.T., J.N., B.L., L.Z., C.V., A.C, R.C., J.P.G., B.C., N.L., C.C., J.A., P.O., and T.B; writing—review and editing, M.T.M.-Q., R.H., J.T., J.N., B.L., L.Z., C.V., A.C, R.C., J.P.G., B.C., N.L., C.C., J.A., P.O., and T.B; supervision, M.T.M.-Q.; funding acquisition and project administration: M.T.M.-Q. All authors have read and agreed to the published version of the manuscript.

## Funding

Funding for this study was provided by FONDECYT N°1240899, awarded by the Agencia Nacional de Investigación y Desarrollo (ANID) of the Chilean Government

## Institutional Review Board Statement

Not applicable.

## Informed Consent Statement

Not applicable.

## Declaration of competing interest

The authors declare that they have no known competing financial interests or personal relationships that could have appeared to influence the work reported in this paper.

## Protocol registration

OFS Home https://osf.io/qjv6m/

## Notes

### Competing Interest Statement

The authors have declared no competing interest.

### Funding Statement

Funding for this study was provided by FONDECYT 1240899, awarded by the Agencia Nacional de Investigacion y Desarrollo (ANID) of the Chilean Government

